# Glaucoma patient screening from online retinal fundus images via Artificial Intelligence

**DOI:** 10.1101/2021.02.11.21251193

**Authors:** Pablo Franco, David Coronado-Gutierrez, Carlos Lopez, Xavier P. Burgos-Artizzu

## Abstract

**Objectives:** To design and evaluate a novel automated glaucoma classifier from retinal fundus images.

**Methods:** We designed a novel Artificial Intelligence (AI) automated tool to detect glaucoma from retinal fundus images. We then downloaded publicly available retinal fundus image datasets containing healthy patients and images with verified glaucoma labels. Two thirds of the images were used to train the classifier. The remaining third of the images was used to create several cross-validation evaluation sets with a realistic glaucoma prevalence, to evaluate the classifier’s performance in a screening scenario.

**Results:** 10,658 retinal fundus images from seven different sources were found and downloaded. They were randomly divided into 7,106 for training and 3,551 for validation. Glaucoma prevalence was 24%. Using the validation set, we created 50 random sets of 1,000 images with a 5% glaucoma prevalence. On these sets, the classifier reached a detection rate of 84.1% (CI 95=+-1.1%) and 95.8% (CI 95=+-0.2%) specificity.

**Conclusions:** A novel glaucoma classifier from retinal fundus images showed promising results as screening tool on a large cohort of patients. A large clinical study is needed to confirm these results.

## Intro

Glaucoma is a neurodegenerative condition that affects the eye, related to increased intraocular pressure^1^. When left untreated, patients may experience a gradual loss of the visual field and can even lose their sight completely^2^. Although there exist many types of glaucoma, all are characterized by damage to the optic nerve that can eventually lead to blindness^3^.

In fact, glaucoma is the second leading cause of blindness worldwide after the cataract, as it affects more than 70 million people^4^. Furthermore, blindness caused by glaucoma is irreversible unlike that caused from cataracts^5^. For this reason, early detection of glaucoma is essential.

Glaucoma can be diagnosed using different techniques. One of the most common is tonometry. This test is done using a device called a tonometer that measures the pressure inside the eye, called intraocular pressure (IOP). The normal value range for IOP is 12-22 mm. Most cases that exceed 22mm are diagnosed with glaucoma ^6^. However, some people may have glaucoma at pressures between 12- and 22-mm Hg. IOP is unique to each person, so this method is controversial^7^.

Another common technique is the visual assessment of the optic nerve. The reason is that one of the typical characteristics of glaucoma is that it produces an excavation or depression in the head of the optic nerve^8^. For this visual assessment of the optic nerve two imaging techniques are used: non-invasive retinal fundus images taken by a digital retinal fundus camera and information from a coherence tomography (OCT)^9^.

Retinal fundus imaging is the preferred technique for a first patient screening due to its simplicity, world-wide availability and low cost compared to OCT, which is more costly^10^. Prior studies on automated glaucoma detection from retinal fundus images can be divided into two different categories: 1) Those based on retinal clinical indicators and 2) those using image texture analysis techniques.

The first type of works rely on a (manual or automatic) segmentation of the optic disc followed by measurements such as the cup-disc ratio (CDR), the cup-disc area, the ISNT rule, the rim-disc ratio (RDR) or the NRR area^11 12 13 14^. For example, Wong et al.^15^ and Joshi et al.^16^ presented different methods to measure CDR, Diaz-Pinto et al.^17^ used CDR, ACDR or ISNT indicators or Wong DWK et tal^13^ used CDR and NRR indicators. The main limitation of these methods is that they require a very accurate disc/cup segmentation and that variations on how the disk is segmented can produce significant changes in performance^8^.

The second type of approaches use image texture analysis techniques, searching retinal fundus images for patterns that determine the presence or not of glaucoma. Machine/Deep Learning is often used to learn from the images what patterns reveal the presence of glaucoma. For example, Chen et al.^18^ used an ensemble network named DENet to automatically classify glaucomatous fundus image directly without segmentation. Abbas ^19^ also used CNN to classify images merging 1200 images achieving specificity of 0.98 and sensitivity of 0.84. Also Orlando et al.^20^ experimented with the use of CNNS applying techniques likes vessels extraction, contrast-limited adaptive histogram equalization (CLAHE) or cropping around the optic nerve head (ONH). Other relevant works (using Deep Learning) are Bajwa et al. ^21^, Syna Sreng et al. ^22^ or Chakravarty et al ^23^.Finally, the REFUGE challenge ^24^ for glaucoma assessment from retinal fundus images has attracted the development of many strategies based on AI by several research groups. Even though the results of many of these prior studies are promising and already point towards the potential of AI for glaucoma screening of patients, none of these studies had as main objective the evaluation of AI in a real screening scenario. As shown later in the Methods section in Table 1, the prevalence of glaucoma in the public glaucoma datasets show a high variance and are very far from the reported real world-wide glaucoma prevalence of 3.5% for population aged 40-80 years^4^.

**Table 1.** Statistics of the different datasets used for the study.

| DATASET | Glaucoma | Normal | Total | Glaucoma prevalence | Origin |
| --- | --- | --- | --- | --- | --- |
| LAG | 1711 | 3143 | 4584 | 37% | China |
| ODIR | 326 | 3095 | 3241 | 10% | China |
| HRF | 15 | 15 | 30 | 50% | Germany |
| SJCHOI86-HRF | 101 | 301 | 402 | 25% | -- |
| DRISHTI-GS1 | 70 | 31 | 101 | 69% | India |
| REFUGE | 120 | 1080 | 1200 | 10% | China |
| ORIGA | 168 | 482 | 650 | 26% | Malaysia |
| **TOTAL** | **2511** | **8147** | **10658** | **24%** |  |

The objective of this study is to fill this gap and evaluate a novel glaucoma AI based classifier designed for patient screening. In order to do so, we adjusted test prevalence to a more realistic number according to large studies of glaucoma^4^ and evaluated our novel AI classifier on this test to judge its potential as screening tool.

## Materials and Methods

### Retinal fundus image datasets

Different public datasets for the evaluation of glaucoma from fundus images are available. To perform this retrospective study, we used datasets Refuge^24^, Origa^25^, LAG ^26^, Drishti-GS1^27^, sjchoi86-HRF ^28^, HRF ^29^ and ODIR^30^. Figure 1 shows image examples from these datasets. Description of each dataset is as follows:

**Figure 1.**
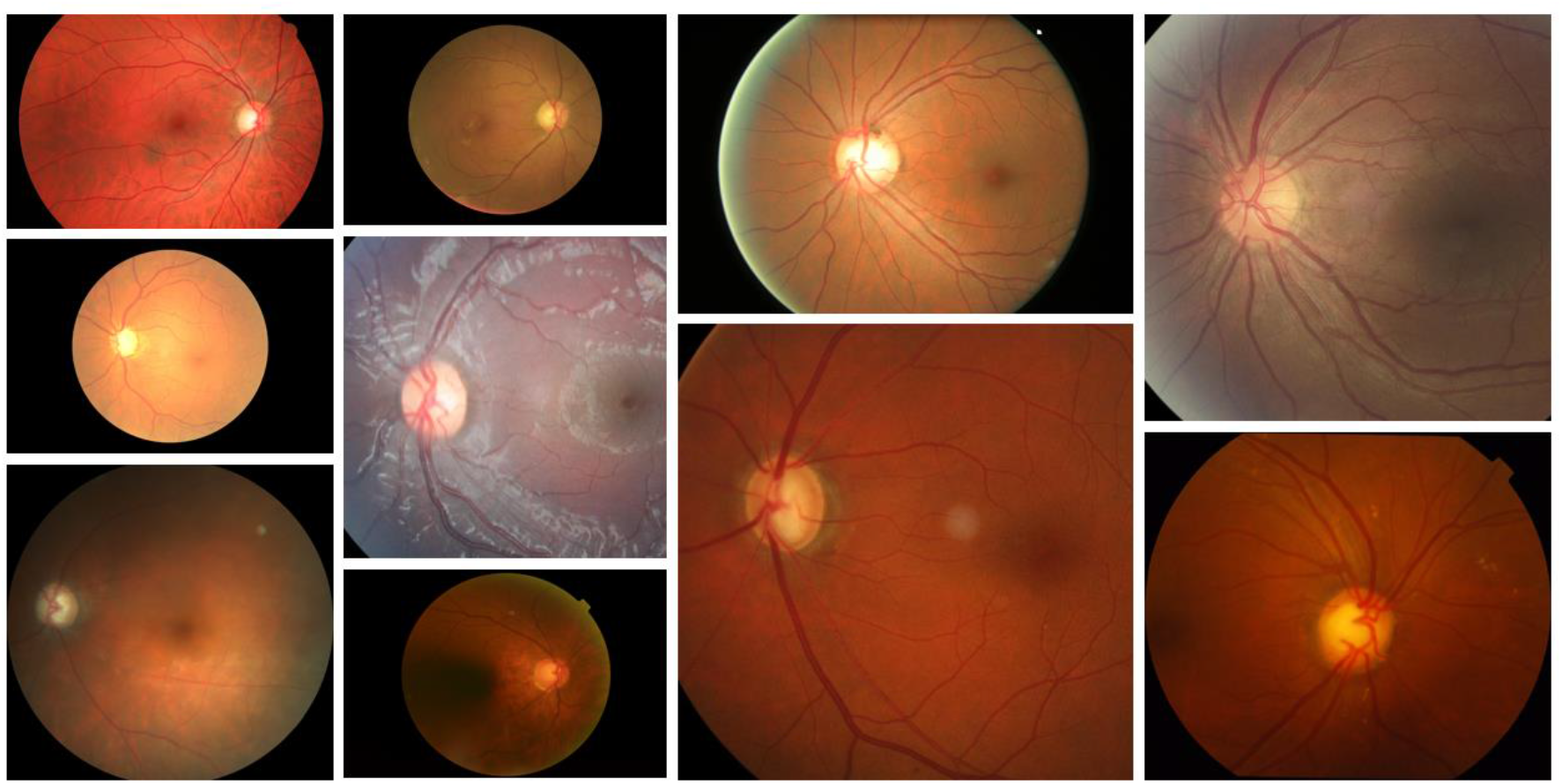
Retinal fundus image examples from the different datasets used.

- **HRF** ^29,31^: This database contains 15 images of healthy patients, 15 images of patients with diabetic retinopathy and 15 images of glaucomatous patients. They were captured by a Canon CR-1 fundus camera with a field of view of 45 grades with a resolution of 3504×2336 px. Binary gold standard vessel segmentation images generated by clinicians are available for each image.
- **LAG** ^26^: This database contains 4,584 fundus images with 1,711 positive and 3,143 negative glaucoma samples obtained from Beijing Tongren Hospital. Each fundus image is diagnosed by qualified glaucoma specialists, taking the consideration of both morphologic and functional analysis, i.e, intra-ocular pressure, visual field loss and manual optic disc assessment.
- **ODIR** ^30^: This database contains images from 5000 patients collected by Shanggong Medical Technology Co., Ltd. from different hospitals/medical centres in China. In these institutions, fundus images are captured by various cameras in the market, such as Canon, Zeiss and Kowa, resulting into varied image resolutions. hey classify patient into eight labels including normal (N), diabetes (D), glaucoma (G), cataract (C), AMD (A), hypertension (H), myopia (M) and other diseases/abnormalities (O) based on both eye images. Glaucoma patient age ranged between 24 and 91 years old.
- **sjchoi86-HRF** ^28^: This dataset contains 601 fundus images divided into 4 groups: normal (300 images), glaucoma (101 images), cataract (100 images) and retina disease (100 images).
- **REFUGE**^24^: This dataset includes 1200 fundus images with ground truth segmentations and clinical glaucoma labels. They were acquired with two different fundus cameras: Zeiss Visucam 500 (2124×2056 pixels) and Canon CR-2 (1634×1634 pixels).
- **ORIGA**(-light)^25^: This dataset contains 650 retinal images collected during the Singapore Malay Eye Study (SiMES) population based study, conducted over a 3 year period on adults aged 40 to 80 years. All labels were annotated by trained professionals from Singapore Eye Research Institute.
- **Drishti-GS1**^**27**^: this dataset contains 101 retinal images. These images were collected at Aravind eye hospital (India) which correspond to patients between 40-80 years of age with a roughly equal number of males and females. Images have been labelled by 4 clinical experts.

In each dataset, we downloaded only normal and glaucoma images. Table 1 summarizes the main statistics of each resulting dataset. These datasets have a large variance in terms of glaucoma prevalence, ranging from 10% to 69%. Considering that global glaucoma prevalence for patients from 40 to 80 years is reported at 3.5% ^4^, not only there is a high variance, but all datasets are overestimating the prevalence of glaucoma. Reporting results on these datasets, as most prior studies do, makes difficult to judge the potential of the proposed tools for screening.

To evaluate the potential of the methods proposed for glaucoma screening, while still using data from all these datasets, we designed a specific testing framework which uses a prevalence close to the 3.5% prevalence reported in large studies in population aged 40-80 years^4^. In order to do so, we first merged all datasets, and then partitioned the images into a training set (66%) and a validation set (33%) keeping the prevalence of each original dataset. Then, for the validation set, for each dataset we created 50 random subsets of 1000 images, each containing 5% glaucoma cases. These sets were used to report average performance and confidence intervals.

### Image pre-processing

As stated in the Intro, in theory, glaucoma affects mainly the optic disc and the area near it. Therefore, most prior works suggest that only the optic disc part of the image should be used. However, we found that this part of the image can be very small in many images and convey very little signal when the image contrast is not set correctly. We explored different alternatives, to establish what part of the image is better suited to detect glaucoma and found that using the full image reached best results. Please see Supp Figure 1 and Supp Table 1 for more info on these experiments. The only preprocessing performed consists in removing the black borders from the input image, see Supp Figure 1.

### Automatic AI glaucoma classifier

A novel glaucoma classifier was developed to classify glaucoma from retinal fundus images. The classifier is a single Deep Convolutional Neural Network^32^ applied on the entire image (after pre-processing to remove the uninformative black borders, see Supp Figure 1). During the development stage, several possible CNN architectures were benchmarked using our AI online platform. The final chosen model was an adapted version of a ResNet-50 architecture^33^.

We relied on Transfer Learning, initially training the CNN on large datasets such as ImageNet Large Scale Visual Recognition Challenge^60^ and Microsoft coco^34^. Then, the net was adapted to the binary classification task and fully retrained (allowing changes to the entire network) using 66% of all the images downloaded. More specifically, 7,106 images were used for training, with a 24% glaucoma prevalence (1,706 images with glaucoma, 5,400 normal). After training, the net was deployed and applied to all the validation images, outputting full probability score (softmax) for glaucoma.

### CNN implementation details

The network wax trained using Transmural Biotech’s AI online platform. The ResNet50 classifier was trained using softmax cross-entropy loss and adam optimizer. 10% of the training set was used as validation set. The first training using ImageNet and Coco images was performed for 90 epochs with a 5-epoch warm-up. Then, the CNN was trained on the training data for a maximum of 15 epochs, early stopping if loss on validation set was not improved for 5 consecutive epochs. Learning rate was adjusted using a cosine decay starting at 1e-4. Batch size was 64 and weight decay was 0.9.

Data augmentation techniques were used during training to improve the classifier robustness and avoid overfitting. Validation images were processed without any alteration nor augmentation tricks.

### Statistical analysis

Statistical analysis was performed using Matlab (Mathworks, USA). The final classifier, once trained, was applied to all validation available images, storing the output glaucoma probability score. The scores were used to draw Receiver Operator Characteristic (ROC) curves and compute full Area Under the Curve (AUC) Then, the ROC curves were used to establish the optimal cutoff points in terms of accuracy. Detection rate, false positive rate (FPR), positive and negative predictive values (PPV and NPV) and positive and negative likelihood ratios (LR+ and LR-) were calculated. We repeated this for each cross-validation set, reporting both the average and 95% Confidence Intervals on the sets.

## Results

Figure 1 shows the ROC curve for glaucoma detection on the 50 cross-validation sets using a total of 3,551 images. AUC was 91.1% +-0.5% and accuracy was 95.3% +-0.2. Metric scores for a cut-off maximizing accuracy are shown in Table 2. The detector achieved a 84.1% detection rate at 95.8%+-0.2% Specificity. Negative Predictive Value was 99.1%+-0.1%.

**Figure 1.**
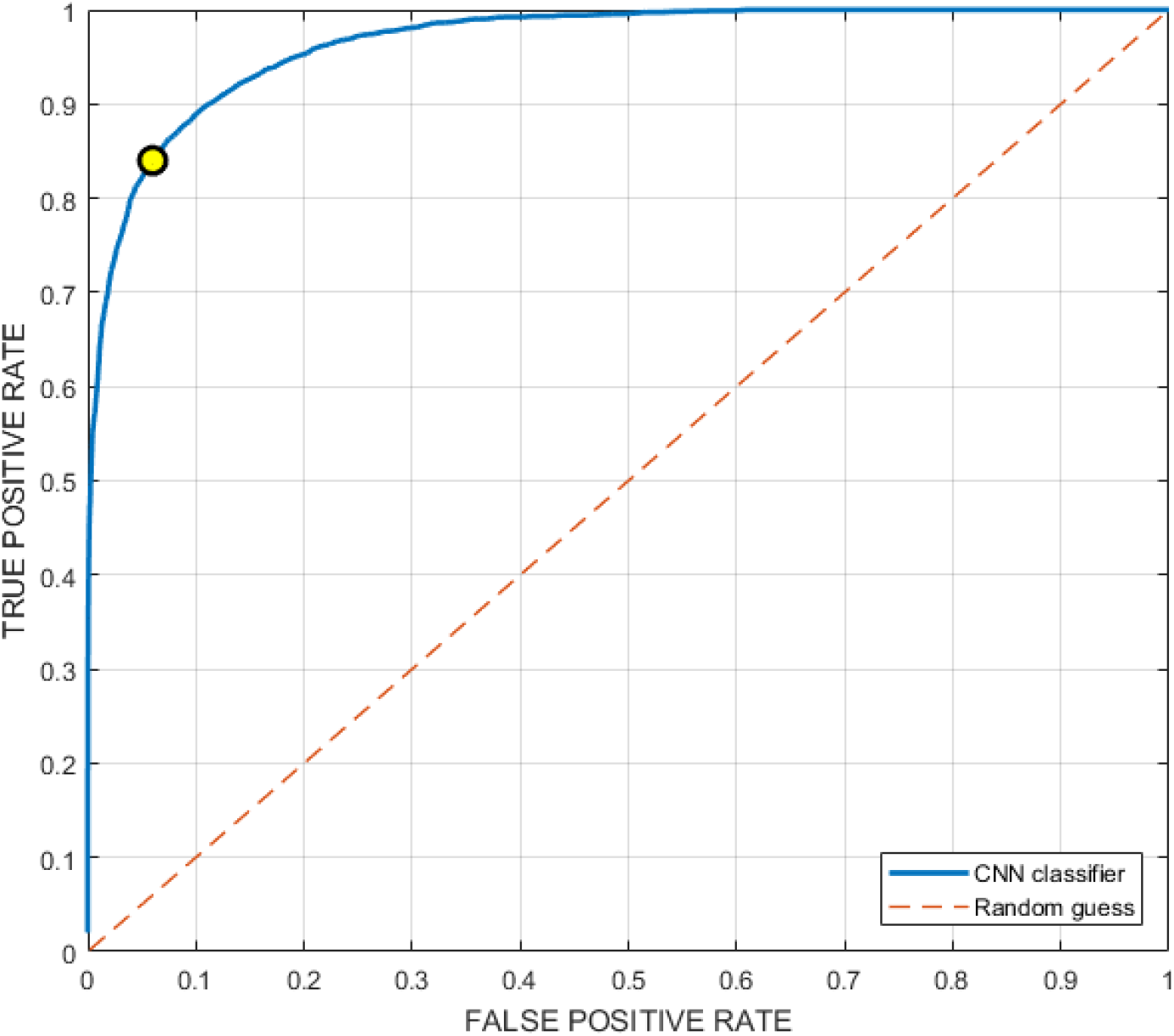
ROC curve for prediction task. The yellow circle marks the optimum cut-off point in terms of accuracy.

**Table 2.** **Metric scores. Values shown as Mean(+- CI 95). ACC=Accuracy. SENS=Sensitivity. SPEC=Specificity**. **PPV=Positive Predictive Value. NPV=Negative Predictive Value. AUC=Area Under the (ROC) Curve**.

| Method | ACC | SENS | SPEC | PPV | NPV | AUC |
| --- | --- | --- | --- | --- | --- | --- |
| CNN | 95.3 %<br>($\pm 0.2$ %) | 84.1 %<br>($\pm 1.1$ %) | 95.8 %<br>($\pm 0.2$ %) | 52.4 %<br>($\pm 1.1$ %) | 99.1 %<br>($\pm 0.1$ %) | 91.1%<br>($\pm 0.5$ %) |

This scenario would have implied, for a hypothetical population of 1000 people with a prevalence of 5% (50 persons with glaucoma), that of the 50 patients which have glaucoma the CNN would have successfully detected 42, leaving 8 undetected. On the other hand, it would have falsely flagged 39/950 patients without glaucoma disease as having glaucoma.

For completeness, supp Figure 2 and Supp Table 2 show individual results on each dataset. Furthermore, although a direct comparison on same data was not possible, Supp Table 3 shows a comparison of our results with respect with prior reported results on some of the datasets. Our novel glaucoma classifier reached similar or superior performance compared against prior methods.

Figure 2 shows example correct glaucoma detections, with the class-activation maps^35^ of the CNN superimposed on top of the image. The CNN appears to be correctly detecting as region of interest the surroundings of optic disc marked in red colour.

**Figure 2.**
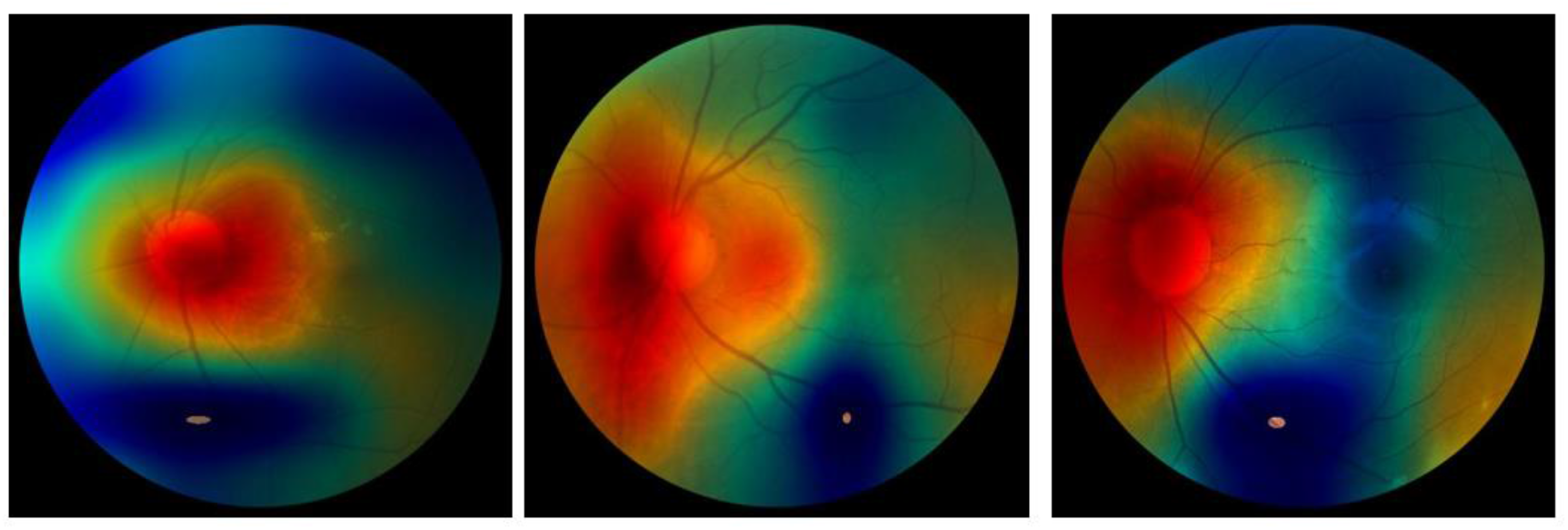
Correct glaucoma detection examples. Original Images are superimposed with the CNN’s Class-activation maps_35_ (red= hot, blue=cold), which provide an estimate of what parts of the image is the CNN model focusing on for classifying the images as glaucoma.

## Discussion

### Main findings

We designed a novel glaucoma classifier based on fully automated analysis of retinal fundus images. The classifier, using state-of-the-art Deep Learning technology, was designed with a clear focus on screening. Tested on thousands of images from seven different sources, the proposed model was capable of detecting 84.1% of the cases at low (0.9%) false positive rates. The classifier performance showed a low variance (5% in terms of sensitivity) even though the quality, dimension and compression of the images used was varied, as shown by Figure 1.

Negative Predictive Value was 99.1%, making it ideally suited to be used in a screening scenario. Results of this study are promising and suggest that the deep learning-based algorithm can detect glaucoma with high accuracy from retinal fundus images, which are easier to obtain in a large population. This highlights its potential application for glaucoma screening.

Compared to reported results from a varied set of prior approaches, our novel glaucoma classifier showed state-of-the-art performance, always reaching similar or superior performance on all the seven datasets tested, which showed a wide variety in resolution, format, contrast and quality of retinal fundus images. The main advantage of our approach is that it does not require a manual optic disc segmentation and is capable of exploiting information from the entire image. A full analysis of this comparison is shown in Supp Table 3.

### Clinical implications

In most cases, a single measurement does not provide enough information for a correct diagnosis of glaucoma^36^. Therefore, traditional glaucoma screening tools typically consider various aspects in combination. During a regular ophthalmological examination, various tests are performed, such as a visual acuity checking, an intraocular pressure measurement or a preliminary analysis of the cup / disc ratio to detect suspicions of the disease. These tests are integrated into different algorithms that, based on the results, will determine if a further detailed evaluation is necessary^37^.

Several studies have evaluated the effectiveness of current glaucoma screening tools. R Khandeka et al.^38^ carried out a study on 3324 patients using ocular pressure and visual inspection of retinal fundus changes as parameters. Using only ocular pressure, the reported sensitivity was 49.7% at 95.6% specificity. Using only disc changes, sensitivity was 48.4% at 97.9% specificity. Sensitivity improved to 67.3% at 96.5% specificity when combining both. They concluded that these parameters were not accurate enough for a trustworthy glaucoma screening. Also in 2013 S-Farzad Mohammadi et al.^39^ evaluated a glaucoma screening algorithm which included examination of the optic disc for vertical cupping, asymmetry, tonometry and automated perimetry over a population of 124 patients (prevalence 4.8%). They reported a Positive Predictive Value of 30%.

Other studies have analysed the performance of screening using other techniques as the Heidelberg Retinal Tomography [HRT]. Healey et al. ^40^ reported 46% of sensitivity and 91% of specificity, Saito et al.^41^ reported a sensitivity of 39% and 96% of specificity. The results from these 2 studies predict that a screening program of 10 000 individuals from a general population with a 2% prevalence of OAG would detect only 80–90 out of the 200 affected, and falsely detect glaucoma on 390–880 normal individuals.

In conclusion, results of the different studios suggest that screening to detect all individuals with glaucoma remains an unsolved challenge^42^. In this context, the method proposed in this study, which is not invasive, automated, capable of providing results in seconds and achieving a sensitivity of 84.1% at 95.8% specificity, could, individually or as an additional step in the screening algorithm, add additional information in order to obtain a more accurate early glaucoma diagnosis.

The proposed methods will be integrated into an online platform, making them available to any professional.

### Strengths and limitations

This study has several strengths. We designed a novel glaucoma classifier fully automated, which is capable to extract information from the full retinal fundus image. The novel classifier was evaluated on a large set of retinal fundus images online which were collected from different centers, showing a wide variety in terms of image resolution, format and quality, mimicking a real clinical scenario. The classifier showed state-of-the-art results when compared directly with prior reported results from a large variety of studies. Furthermore, we combined combine all available retinal fundus image datasets while adjusting the prevalence of glaucoma in order to evaluate our classifier in a scenario realistic from a clinical point of view. Finally, to further imitate realistic conditions, we did not filter the images by imposing our own quality constraints and used all online images available directly.

We acknowledge two main limitations. Firstly, the study was conducted on online images and therefore we weren’t able to neither double-check clinical outcomes of the images nor verify the role image acquisition had on performance (a known Deep Learning issue), since the specific image acquisition details from each source was not provided. Secondly, each source had varied glaucoma prevalence and even though we tried to mitigate this issue by adjusting prevalence manually and using images acquired by several different centers, these results cannot substitute a proper, large clinical validation study, which should and will be pursued.

As future work, information provided by AI methods could be combined with all other information about the patient (presence of a positive family history, medical history, ethnicity, age, gender). This information was not available in the public datasets used. Additional evaluation using these variables in order to detect relations would be very interesting.

## Conclusions

A novel glaucoma classifier from retinal fundus images showed promising results as screening tool on a large cohort of patients. The new tool could help in the first screening of patients, improving screening accuracy compared to visual inspection by the oftalmologists. A large clinical study is needed to confirm these results.

## Data Availability

All used data is fully available since it is from public and open-source online datasets.

## Data and materials availability

All data associated with this study is available in the main text and comes from public datasets. The developed glaucoma detection method will be made available through a novel platform.

