## Supplementary material for "Glaucoma patient screening from online retinal fundus images via Artificial Intelligence"

**Supplementary material for paper** Glaucoma patient screening from retinal fundus images via Artificial Intelligence

Transmural Biotech S. L. Barcelona, Spain.

### *Best ROI for image processing*

Using the dataset REFUGE, which included the ground truth optic disc segmentation for 1200 images, we explored what region of the image was the more indicated for glaucoma detection. We tested 3 regions, shown in Supp Figure 1: a) the full fundus image, b) the optic disc, c) a bounding box containing the optic disc enlarged 1.5 times. Note that for the first ROI (full image), we applied an automatic process to detect and remove the black borders. The other two were computed from the manual optic disc ROI provided.

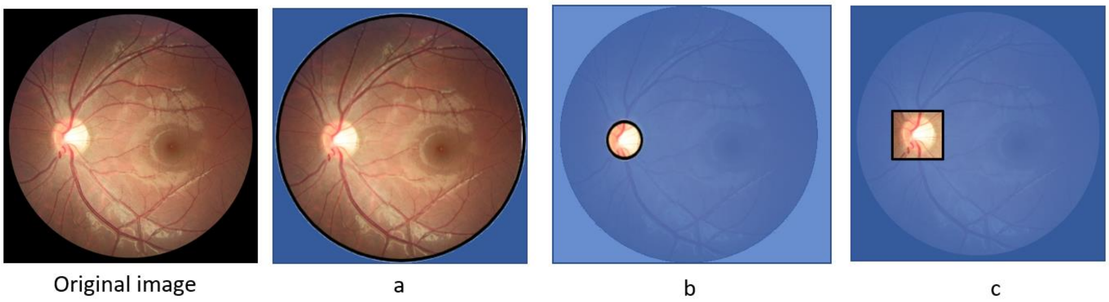


**Supp Figure 1: Examples of ROIs considered. a) Full image ROI (automatic), b) optic disc ROI (manual), c) Enlarged bounding box around optic disc (manual).**

For each one of the three ROIs, we trained a CNN classifier using the 800 training images from REFUGE and we evaluated its performance on the 400 test images. Results are shown in Supp Table 1. The three achieved very similar results with differences of less than 1% in Specificity at identical Sensitivity. Full image achieved an Accuracy 97.5 and a F1 score of 86.1 while optic disc only achieved an Accuracy of 96.8 and a F1 Score of 82.7 and bounding box enlarged achieved an Accuracy of 97 and a F1 score of 83.8. We chose to use the full image for our glaucoma detector due to the slightly better results and its automated nature.

| | FULL IMAGE | | | | OPTIC DISC | | | | BOUNDING BOX | | | |
|---|---|---|---|---|---|---|---|---|---|---|---|---|
| | ACC | SENS | SPEC | F1 | ACC | SENS | SPEC | F1 | ACC | SENS | SPEC | F1 |
| CNN | 97.5 | 77.5 | 99.7 | **86.1** | 96.8 | 77.5 | 98.9 | 82.7 | 97 | 77.5 | 99.2 | 83.8 |

**Supp Table 1. Comparative of glaucoma detection using the three ROIs on REFUGE dataset.**

*Results by dataset*

As discussed in the paper and shown in Table 1, the datasets evaluated have a large variance in terms of glaucoma prevalence and quality of images. Most datasets have a very unrealistic glaucoma prevalence which makes difficult the analysis and validation of a real-world clinical glaucoma screening tool. In the paper we fixed the prevalence of our test set to 5% and used random samples from each dataset with that fixed prevalence.

However, for completeness, we report here the results achieved by our tool in each one of these datasets separately without altering the test set's prevalence. Supp Figure 2 shows the ROC curves, while Supp Table 2 shows the individual results after choosing a cut-off maximizing accuracy.

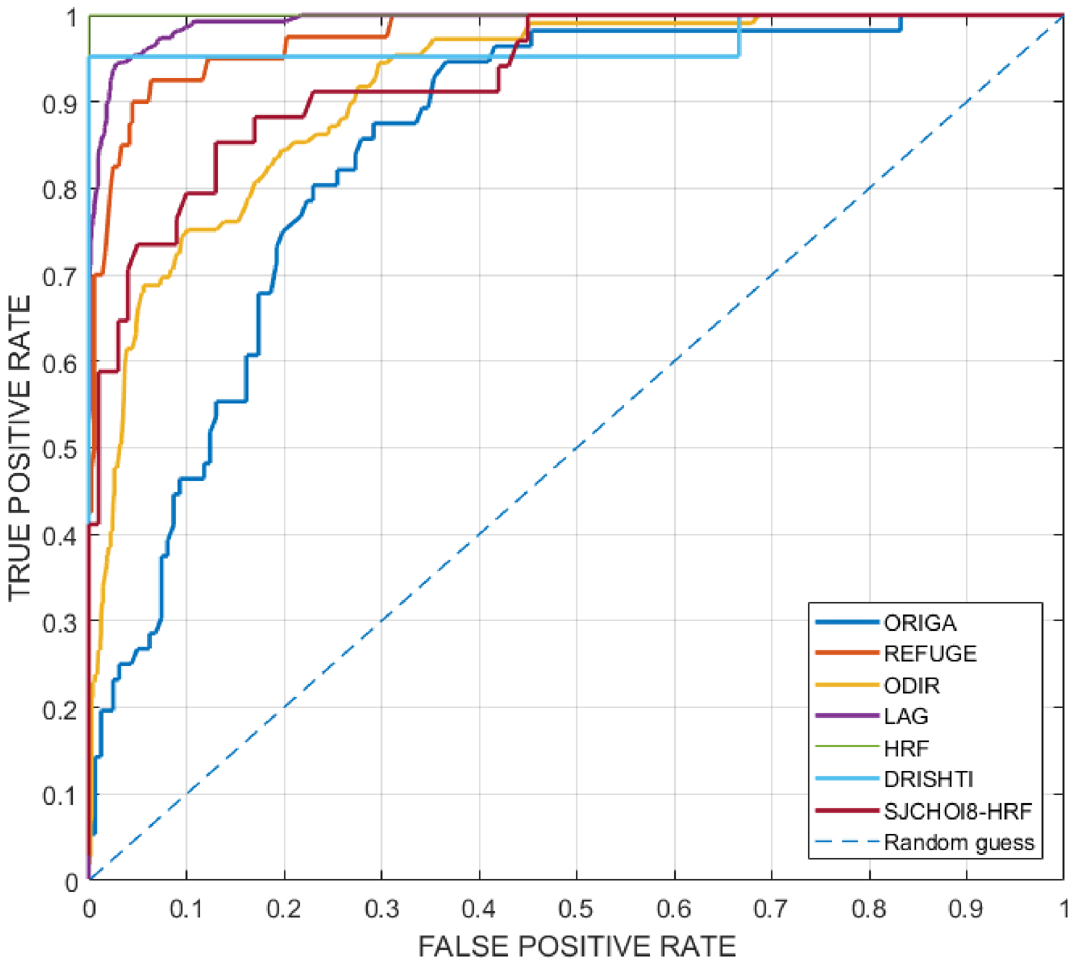


**Supp Figure 2. ROC Curves for each dataset**

| Method | Acc. | F1 | AUC | Detection Rate | False Positive Rate |
|---|---|---|---|---|---|
| **LAG** | 1501/1619 (92.7 % +-0.1 %) | 90.6 % +-0.1 % | 99.0 % +-0.0 % | 567/571 (99.3 % +-0.1 %) | 114/1048 (10.9 % +-0.1 %) |
| **ODIR** | 972/1141 (85.2 % +-0.7 %) | 49.6 % +-1.3 % | 84.3 % +-0.4 % | 83/109 (76.1 % +-0.7 %) | 143/1032 (13.9 % +-0.8 %) |
| **ORIGA** | 171/217 (78.8 % +-0.6 %) | 57.4 % +-1.5 % | 71.3 % +-1.0 % | 31/56 (55.4 % +-1.9 %) | 21/161 (13.0 % +-0.5 %) |
| **REFUGE** | 354/400 (88.5 % +-1.1 %) | 62.3 % +-2.5 % | 96.0 % +-0.4 % | 38/40 (95.0 % +-0.7 %) | 44/360 (12.2 % +-1.2 %) |
| **HRF** | 10/10 (100.0 % +-0.0 %) | 100.0 % +-0.0 % | 100.0 % +-0.0 % | 5/5 (100.0 % +-0.0 %) | 0/5 (0.0 % +-0.0 %) |
| **DRISHTI-GS1** | 29/30 (96.7 % +-0.7 %) | 97.6 % +-0.5 % | 97.4 % +-0.6 % | 20/21 (95.2 % +-1.0 %) | 0/9 (0.0 % +-0.0 %) |
| **SJCHOI86-HRF** | 116/134 (86.6 % +-0.7 %) | 76.3 % +-1.2 % | 89.5 % +-0.8 % | 29/34 (85.3 % +-1.4 %) | 13/100 (13.0 % +-0.7 %) |

**Supp Table 2. Metric scores on each dataset**

As expected, due to the high variance of glaucoma prevalence present in each dataset (see Table 1 in our paper) results show important variations in the performance depending on the dataset. For example, fixing as reference max false positive rate of 15%, for the dataset HRF our ACC is 100% but in case of ORIGA is 78.8%. This situation is very common with the public datasets available for different reasons such as the quality of images, criteria for labelling or the origin of the population sample, as reported previously in other studies [1] [2] [3].

*Comparison with prior approaches*

As far as we know, we are the first to combine all available datasets together and take into account the prevalence of glaucoma in general population in order to develop and evaluate a glaucoma screening tool targeting its real clinical use. We cannot directly compare our tool with prior approaches on the same data, but we can compare the results obtained by prior methods as reported by its authors on the test set of some of the datasets we used. However, this is still a partially incorrect comparison since many other differences remain. To name just a few: the number of images from the dataset used to train and test the models varies across studies; Some studies apply a manual filtering of images according to their quality; Some studies train models using only images from the same dataset while others combine images from several other datasets, or even use their own private images during training. Nevertheless, with this in mind, we show this comparison in Supp Table 3.

| Dataset | ACC | AUC | Reference | Annotations |
|---|---|---|---|---|
| LAG | 95.3 | 97.5 | Li, Liu & Xu [4] | AG-CNN 2 stages network architecture: pathological localization CNN and CNN for glaucoma classification. |
| | 80.2 | 95.6 | Li, Liu & Xu [4] | Using Chen et al. deep learning architecture[3] based on six layers CNN , performance referenced in paper. |
| | 89.7 | 90.1 | Li, Liu & Xu [4] | Using Li et al. architecture in paper, performance referenced in paper. |
| | **92.7** | **99** | **Ours** | |
| SJCHOI86-HRF | 70.82 | 77.39 | Diaz-Pinto et al. [2] | CNN Xception architecture using a bounding box around optic disc as ROI. |
| | **86.6** | **89.5** | **Ours** | |
| HRF | 80.0 | 83.5 | Diaz-Pinto et al. [2] | CNN Xception architecture using a bounding box around optic disc as ROI. |
| | 86.7 | 89 | Sidong Liu et al. [5] | CNN ResNet50 architecture using HRF images just for testing, not for train. |
| | **100** | **100** | **Ours** | |
| ODIR | Not comparisons found. | | | |
| Drishti-GS1 | 75.2 | 80.41 | Diaz-Pinto et al. [2] | CNN Xception architecture using a bounding box around optic disc as ROI. |
| | 76.7 | 78 | Chakravarty et al.[6] | Deep Learning method trained with private dataset of 386 images and tested against DRISHTI-GS1 dataset. |
| | 75.2 | 80.4 | Orlando et al.[7] | CNN method trained and tested only with splits of Drishti dataset. |
| | 90 | 92.0 | Syna Sreng et al. [1] | Ensemble of CNN methods trained and tested only with splits of DRISHTI-GS1 dataset. |
| | **96.7** | **97.4** | **Ours** | |
| REFUGE | 86.3 | 96.4 | Winner of refuge Challenge CHMUK Team [8] | CDR values computed from ellipses fitted to automated OD/OC segmentations |
| | 85.2 | 93.8 | Chen et al. [9] | Segmentation and use of CDR Ratio measured manually. |
| | 83.8 | 88.6 | Chen et al. [9] | Segmentation and use of CDR Ratio measured automatically. |
| | 82.2 | 82.01 | Chen et al. [9] | Ensemble DL method DeNet using whole fundus image and disc optic segmentation trained and tested only with splits of REFUGE dataset. |
| | 95.5 | 95.1 | Syna Sreng et al. [1] | Ensemble of CNN methods trained and tested only with splits of REFUGE dataset. |
| | **88.5** | **96** | **Ours** | |
| ORIGA | 83.5 | 88.8 | Syna Sreng et al. [1] | Ensemble of CNN methods trained and tested only with splits of ORIGA dataset. |
| | 76.9 | 83.1 | GUO et al. [10] | Random Forest-based glaucoma classifier trained and tested only with splits of ORIGA dataset. (CDR, ISNT) with Disc optic segmentation. |
| | 79.67 | 86.8 | Bajwa et al. [11] | CNN methods trained and tested only with splits of ORIGA dataset. |
| | **78.8** | **71.3** | **Ours** | |

**Supp Table 3. Comparison with similar works**

**LAG** : Li, Liu & Xu et al. [4] performed three experiments using different methods: a) using their own method with AG-CNN networks, reporting ACC=95.3 and AUC=97.5, b) using the Chen et al. architecture[3], reporting ACC=80.2 and AUC=95.6 and c) using Li et al. architecture[4] , reporting ACC=89.7 and AUC=90.1. Our method achieved comparable results reaching ACC=92.7 and AUC=99.

**REFUGE:** is the dataset used for the Retinal Fundus Glaucoma challenge[8], and therefore there are many results reported. We have chosen some of the most representative according to the different techniques used for the analysis (CNN, manual CDR measurement or automatic CNN measurement). In this case our selection for the comparison was the last winner of the REFUGE challenge[8] and the three results reported by Chen et al[9] using diverse approaches. Fixing the specificity at 85% CHMUK Team[8], winner of the REFUGE challenge 2018 reported ACC=86 and AUC=96.44. Chen et al[9] reported: a) ACC=85.2 and AUC=93.86 with manual CDR from trained clinician, b) ACC=83.8 and AUC=88.65 using automatic CDR scores, and c) ACC=82.2 and AUC=82.01 using CNN scores. Our metric scores were ACC=88.5, AUC=96, therefore comparable to the best previously reported scores.

**SJCHOI86-HRF:** was used by Diaz-Pinto et al. [2] to train a CNN Xception architecture. They reported an ACC=70.82 and AUC=77.39. Our results were ACC=86.6 and AUC= 89.5 clearly improving their performance.

**Drishti-GS1**: we compare against a) Diaz-Pinto et al. [2] using a CNN Xception architecture, which achieved ACC: 75.25 and AUC 80.41, b) Chakravarty et al.[6] which achieved ACC=76.7 and AUC=78 using deep Learning method trained with a private dataset c) Orlando et al.[7] which achieved ACC=75.2 and AUC=80.4. and d) Syna Sreng et al.[1] which achieved the best performance with ACC=90 and AUC=92.06 using an ensemble of CNN. Our scores were ACC=96.7 and AUC=97.4, improving the cited references.

**ORIGA**: dataset scores achieved were ACC=78.8 and AUC=71.3 while the best results of Syna Sreng et al.[1] using an ensemble of CNN methods were ACC=83.5 and AUC=88.8. This dataset also was used by GUO et al.[10] applying clinical indicators like CDR, score achieved were ACC=76.9 and AUC=83.1. Finally, Bajwa et al. [11] using CNN achieved ACC=79.67 and AUC=86.8. In this case our results were a little worse probably because they were training only with images from the same dataset that was tested.

**HRF**: dataset best scores reported by Diaz-Pinto et al.[2] using a CNN Xception architecture were ACC=80 and AUC=83.54 and scores of Sidong Liu et al. [5] using a ResNet50 architecture achieved ACC=86.7 and AUC=89. Our scores where ACC=100 and AUC=100. This dataset is very small and only 10 images were included in the testing split, and therefore is not very statistically significant.

**ODIR**: dataset, we have not found any prior studies reporting results.

In conclusion, after comparing the results of our method with prior proposed methods on seven different databases, it is clear that our method achieves state-of-the-art results.
